# Associations between attention-deficit hyperactivity disorder genetic liability and ICD-10 medical conditions in adults: Utilizing electronic health records in a Phenome-Wide Association Study

**DOI:** 10.1101/2022.11.28.22282824

**Authors:** Elis Haan, Kristi Krebs, Urmo Võsa, Isabell Brikell, Henrik Larsson, Estonian Biobank Research Team, Kelli Lehto

## Abstract

**Background:** Attention-deficit hyperactivity disorder (ADHD) is often comorbid with other medical conditions in adult patients. However, ADHD is extremely underdiagnosed in adults and little is known about the medical comorbidities in undiagnosed adult individuals with high ADHD liability. In this study we investigated associations between ADHD genetic liability and electronic health record (EHR)-based ICD-10 diagnoses across all diagnostic categories, in individuals without ADHD diagnosis history.

**Methods:** We used data from the Estonian Biobank cohort (N=111,261) and generated polygenic risk scores (PRS) for ADHD (PRS_ADHD_) based on the ADHD genome-wide association study. We performed a phenome-wide association study (PheWAS) to test for associations between standardized PRS_ADHD_ and 1,515 EHR-based ICD-10 diagnoses in the full and sex-stratified sample. We compared the observed significant ICD-10 associations to associations with: 1) ADHD diagnosis and 2) questionnaire-based high ADHD risk analyses.

**Results:** After Bonferroni correction (p=3.3×10^-5^) we identified 80 medical conditions associated with PRS_ADHD_. The strongest evidence was seen with chronic obstructive pulmonary disease (OR=1.15, CI=1.11-1.18), obesity (OR=1.13, CI=1.11-1.15), and type 2 diabetes (OR=1.11, CI=1.09-1.14). Sex-stratified analysis generally showed similar associations in males and females. Out of all identified associations, 40% and 78% were also observed using ADHD diagnosis or questionnaire-based ADHD, respectively, as the predictor.

**Conclusions:** Overall our findings indicate that ADHD genetic liability is associated with an increased risk of a substantial number of medical conditions in undiagnosed individuals. These results highlight the need for timely detection and improved management of ADHD symptoms in adults.

## Introduction

Attention-deficit hyperactivity disorder (ADHD) is a highly heritable common neurodevelopmental disorder with a worldwide prevalence of about 5% among school-aged children (Polanczyk, de Lima, Horta, Biederman, & Rohde, 2007). Although the onset of ADHD is typically in childhood, ADHD often persists in adulthood. It has been shown that at least 15% of children with ADHD meet diagnostic criteria for ADHD also in adulthood (Faraone, Biederman, & Mick, 2006). The average prevalence of ADHD in adults has been found to be 3-4% (Fayyad et al., 2007). However, ADHD in adults is often underdiagnosed and/or untreated, which may lead to negative psychosocial and health consequences (Ginsberg, Quintero, Anand, Casillas, & Upadhyaya, 2014). Evidence from previous studies have shown substantial comorbidity between ADHD and both psychiatric and somatic medical conditions, e.g. hypertension, migraine, obesity and type 2 diabetes, in adulthood (Brikell, Burton, Mota, & Martin, 2021; Chen et al., 2018; Kittel-Schneider et al., 2022). Yet, the comorbidities in undiagnosed or subclinical ADHD populations are less clear.

Although shared genetic factors have been implicated in explaining the link between ADHD and psychiatric outcomes (Brikell et al., 2021), little is known about the role of shared genetic mechanisms in ADHD and medical conditions. ADHD is highly heritable with an average heritability of 76% in twin studies (Faraone & Larsson, 2019; Faraone et al., 2005) and the heritability attributed to single nucleotide polymorphisms (SNPs) identified in large genome-wide association studies (GWAS), e.g. SNP-heritability of 22% (Demontis et al., 2019). The genetic correlations based on the results of ADHD GWAS showed significant genetic overlap between ADHD and several mental health (e.g. depression, autism, subjective well-being and neuroticism), as well as physical health traits and conditions (e.g. body mass index (BMI), obesity, HDL cholesterol and type 2 diabetes) (Demontis et al., 2023, 2019). Another approach to investigate shared genetics across traits and disorders is by using polygenic risk scores (PRSs), which capture an individuals weighted sums of risk alleles as detected with a large-scale, independent GWAS (Lewis & Vassos, 2020). Currently, PRS for ADHD (PRS_ADHD_) based on the GWAS explains up to 5.5% of variance in ADHD and individuals in the top 10% of this PRS show 5 times higher odds of ADHD compared to individuals in the lowest PRS decile (Demontis et al., 2019). Although many individuals with high PRS_ADHD_ will not have ADHD diagnosis, PRS_ADHD_ has still great potential to identify individuals with high ADHD genetic liability in populations with low ADHD prevalence, potentially expressing a higher degree of ADHD like traits (e.g. impulsivity, inattention).

Despite the strong advantage of exploiting electronic health records (EHR) in medical genetics research in biobanks (Abul-Husn & Kenny, 2019) there are currently few available reports on the associations between PRS_ADHD_ and EHR-based medical diagnoses. Such studies would help to generate hypothesis about potential underlying mechanisms, e.g. shared genetic etiology, confounding or causal effects seen in studies of ADHD and somatic health comorbidities. A phenome-wide association study (PheWAS) is a hypothesis-free approach that can be used to test associations between a single predictor (e.g. genetic variants, PRS, diagnosis) and a wide range of phenotypes, such as the EHR data (Pendergrass et al., 2011). A PheWAS based on UK Biobank questionnaire and diagnosis data showed that PRS_ADHD_ was associated with several health related traits (e.g. physical abuse, younger age at first sexual intercourse, smoking behaviour, obesity, higher BMI and several blood measures) (Leppert et al., 2020). Another EHR-based PheWAS in 10,000 Penn Medicine Biobank Cohort participants using more than 1,800 phecodes over 6,6 years on average also found PRS_ADHD_ associations with medical conditions (e.g. tobacco use disorder, chronic airway obstruction, type 2 diabetes) (Kember et al., 2021).

We aimed to take the hypothesis-free PheWAS approach in a large population-based biobank (Estonian Biobank; EstBB) to identify the associations between ADHD genetic liability and lifetime history of medical conditions based on ICD-10 diagnoses from EHRs in individuals without a history of ADHD diagnosis (PRS_ADHD_ analysis). Considering the sex differences in the prevalence of ADHD (Biederman, Faraone, Monuteaux, Bober, & Cadogen, 2004; Gaub & Carlson, 1997), we also investigated potential sex differences for the associations between PRS_ADHD_ and medical conditions. Additionally, we compared the direction and strength of the identified ICD-10 associations that passed multiple correction to the associations between these medical conditions and EHR-based ADHD diagnosis (ADHD diagnosis analysis) as well as to the high ADHD risk based on an adult ADHD brief screening instrument (questionnaire-based ADHD analysis). Although existing evidence shows substantial comorbidity and genetic correlation between ADHD and depression, as well as depression and other medical conditions (Carney & Freedland, 2017; Demontis et al., 2023, 2019; Kan et al., 2016; Katzman, Bilkey, Chokka, Fallu, & Klassen, 2017; Levey et al., 2021; Rajan et al., 2020), it is unknown to what degree depression may mediate ADHD-medical condition link in adults. Therefore, our secondary aim was to explore mediation effect by depression in the identified PRS – medical condition associations.

## Methods and Materials

### Study Population

The Estonian Biobank (EstBB) is a large data-rich population-based biobank (Leitsalu et al., 2015), covering approximately 20% of the adult population in Estonia (N=∼210,000; 66% females; mean birth year 1971). All EstBB participants have signed an informed consent form and provided blood samples for genotyping. The EHR data is regularly retrieved by linking to the national health databases and registries, such as the National Health Insurance Funds (NHIF) database, cause of death register, and hospital records. Estonia has universal health care, covering more than 95% of the population. The research project has obtained approval from the Estonian Council on Bioethics and Human Research. More details about the recruitment of participants can be found in Supplementary information.

### Phenotype data

#### Medical conditions via ICD-10 diagnoses

Medical diagnosis data was drawn from the EHRs based on the NHIF’s database covering the period from 2004 to 2020. NHIF’s database is a nationwide system integrating data from all healthcare providers in Estonia. EHR data included 2,004 ICD-10 codes, of which 489 were excluded as these were not medical diagnoses or were in a very low prevalence (Table S1). In total, 1,515 phenotypes from all ICD-10 disease categories (Table S2) were included in the analyses. ADHD diagnosis was defined by ICD-10 diagnosis code F90 (including diagnoses F90.0, F90.1, F90.8 and F90.9).

#### High ADHD risk based on Adult ADHD Self-Report Scale

As a sensitivity analysis, we included self-reported ADHD symptoms score, measured with the modified Estonian version of Adult ADHD Self-Report Scale (ASRS v1.1) part A (Kessler et al., 2005). ASRS part A is a brief six-item screening questionnaire and items are assessed on Likert scale ranging from 0-4: 0=never; 1=rarely; 2=sometimes; 3=often; 4=very often. A score cutoff of 14 out of 24 represents individuals with higher risk for ADHD (Kessler et al., 2007). ASRS data in EstBB is available for 86,245 participants.

### ADHD polygenic risk scores

To compute the PRS, the PRS-CS-auto algorithm was first used to apply a Bayesian regression framework to infer posterior effect sizes of SNPs by using the summary results from a large Psychiatric Genomics Consortium ADHD GWAS (Demontis et al., 2019) and an external linkage disequilibrium reference panel (Ge, Chen, Ni, Feng, & Smoller, 2019). Per-individual risks were further calculated with plink 2.0 (Chang et al., 2015). The PRS_ADHD_ was standardized for the PheWAS analysis and categorized into deciles for subsequent analyses to test differences in high vs low/middle genetic liability groups. Prior to the main analysis, we tested the association between PRS_ADHD_ and ICD-10 ADHD diagnosis history. The variance explained by the PRS_ADHD_ was computed by subtracting the Nagelkerke pseudo R2 from the full (PRS, birth year, recruitment year, sex and 10 genetic principal components (PCs)) and the reduced (birth year, recruitment year, sex and 10 PCs) logistic regression models. Details about the genotyping and imputation procedures in the EstBB can be found in Supplementary information.

### Statistical analysis

#### Main analysis

PheWAS was implemented to test for the associations between PRS_ADHD_ and 1,515 ICD-10 diagnosis codes in the undiagnosed subsample where all individuals with a history of ADHD diagnosis (F90*) were removed (N=464). Individuals with ADHD diagnosis were excluded because compared to the PRS_ADHD_ analysis sample, they were significantly younger and have likely received treatment or other support to manage their ADHD-related risks. Analyses were performed as described in our pre-registered protocol (Haan, Krebs, Võsa, & Lehto, 2021). Analyses were conducted using logistic regression and adjusted for sex (except in sex-stratified analyses), birth- and recruitment year to adjust for birth cohort effects and different EstBB recruitment strategies used across two decades, and 10 PCs to adjust for genetic ancestry (except in secondary analyses). Relatedness was handled by excluding related individuals separately in the PRS_ADHD_ and ADHD diagnosis analysis samples (PI-HAT>0.2) which remained 111,261 individuals in the PRS_ADHD_ analysis sample and 111,601 individuals in the ADHD diagnosis sample. Additionally, for comparison we ran the main analyses in the full cohort without excluding related individuals (N=196,935). We corrected for multiple testing by applying Bonferroni correction (p-value threshold 3.3×10^-5^). PheWAS and mediation analyses were run in R (version 3.6.0). PheWAS results were visualised using the PheWAS library https://github.com/PheWAS/PheWAS. Other analyses were performed using Stata v.17.

#### PRS_ADHD_ deciles

We further examined the associations that survived the multiple testing correction in the PheWAS by using PRS deciles (ranging from 10^th^ to 90^th^ percentiles) to explore the difference in the odds of being diagnosed with a specific medical condition in high vs low (10^th^ vs 1^st^ PRS decile) and in high vs middle (10th vs 5^th^ PRS decile) ADHD genetic liability groups. While the difference in the top and bottom genetic liability groups reflects the differences between the two extreme ends of ADHD genetic liability, the top versus middle comparison reflects the differences between the highest and the population average ADHD genetic liability groups. Additionally, we explored sex differences in the top and bottom PRS decile analysis.

### Secondary analyses

#### Comparison of effects across PRS_ADHD_, ADHD diagnosis and questionnaire-based ADHD analyses

To shed light on the different patterns in medical condition risk in biobank participants with high ADHD genetic liability but without an ADHD diagnosis, EHR-based ADHD diagnosis and clinically relevant ADHD symptoms, we conducted two sets of additional logistic regression analyses only on these medical outcomes that passed the multiple testing correction in the PRS_ADHD_ PheWAS analysis. First, we used the EHR-based ADHD diagnosis as the predictor and second, we used the questionnaire-based high ADHD risk as the predictor.

#### Causal mediation analysis with depression diagnosis

Considering the well-established link between depression, ADHD and more severe somatic health conditions, we ran causal mediation analysis for the identified phenotypes and included lifetime depressive episode diagnosis as a mediator. Lifetime depressive episode diagnosis was defined based on the ICD-10 diagnosis codes F32* and F33*. Causal mediation analysis was run using the R package ‘mediatioń and 1000 simulations was used for calculating estimates (Tingley, Yamamoto, Hirose, Keele, & Imai, 2014). Causal mediation analysis provides estimates for the average causal mediation effect, average direct effect and total effect. Here we were interested in quantifying the proportion of effects on the medical conditions that were mediated by depression.

## Results

A descriptive overview of the study samples is shown in Table 1. The main analysis study sample (PRS_ADHD_) size was 111,261 individuals, after exclusion of related individuals (N=85,674), ADHD cases (N=464), and individuals with missing EHR records (N=2,887). 65% of the study sample were female and the mean birth year was 1970. In the ADHD diagnosis analysis sample, the sample size was 111,601, of which 464 were diagnosed ADHD cases (0.5%). In the questionnaire-based ADHD sample, 3,556 participants screened positive for high risk for ADHD (8%). Cross-group comparisons indicate that although the female-male ratio was similar in all three samples, individuals in the ADHD diagnosis analysis sample were considerably younger (mean birth year 1990) compared to the two other study samples (mean birth year 1970 for PRS_ADHD_ analysis sample and 1979 for questionnaire-based ADHD analysis sample).

**Table 1.** Descriptive overview of study samples

|  | PRS<sub>ADHD</sub> analysis sample | ADHD diagnosis analysis sample | Questionnaire-based ADHD analysis sample* |
| --- | --- | --- | --- |
|  |  | Cases/controls | Cases/controls |
| **N** | 111,261 | 464 (0.4%)/111,137 (99.6%) | 3,556 (8%)/41,152 (92%) |
| **Sex** |  |  |  |
| **Men** | 38,652 (35%) | 283 (61%)/38,616 (35%) | 917 (26%)/12,293 (30%) |
| **Women** | 72,609 (65%) | 181 (39%)/72,521 (65%) | 2,639 (74%)/28,859 (70%) |
| **Birth year (mean, SD)** | 1970 (16.5) | 1990 (10.3)/1971 (16.5) | 1979 (13.3)/1972 (14.1) |
| **Number of ICD-10 diagnoses (mean)** | 41 | 48/41 | 46/39 |
| **Education\*\*** |  |  |  |
| **Primary** | 7,045 (9%) | 61 (29%)/7,040 (9%) | 198 (7.6%)/1,252(4.2%) |
| **Secondary** | 38,092 (50%) | 109 (52%)/38,261 (50%) | 1,225 (45.4%)/16,629 (42.8%) |
| **Higher** | 31,790 (41%) | 41 (19%)/31,763 (41%) | 1,239 (47%)/17,507 (53%) |
| **Missing** | 34,334 | 253/34,284 | 894/8,426 |
| **Depression diagnosis** | 19,997 (26%) | 98 (46%)***/19,970 (26%) | 1,205 (45%)/7,718 (24%) |
| **ADHD diagnosis** | 0 | 464 | 44 |

Note: \*positive screening; \*\*based on the complete set subset ( $N_{ADHD\ PRS}=76,927$ ; $N_{ADHD\ diag}=211/77,064$ ; $N_{ADHD\ ASRS}=2,662/35,388$ ), education was categorised as primary (1-4 grades, 5-9 grades or no education; secondary (10-12 grades and vocational education); higher (university and research degree); \*\*\*data available for 211 participants in the ADHD diagnosis analysis sample and for 2,662 participants who screened positive for ADHD in the questionnaire-based ADHD analysis sample; PRS – polygenic risk score; SD – standard deviation

### PheWAS of PRS_ADHD_

First, we tested the association of the PRS_ADHD_ with ADHD diagnosis (OR=1.34, CI=1.22-1.47) and questionnaire-based ADHD (OR=1.06, CI=1.02-1.09). Results indicate that 1 SD increase in PRS_ADHD_ corresponds to 34% increase in the odds of ADHD diagnosis and 6% increase in the odds of higher risk for self-reported questionnaire-based ADHD. PRS_ADHD_ explained 0.6% variance in EHR-based ADHD diagnosis and 0.04% in questionnaire-based high-risk ADHD.

Our PheWAS showed evidence of association between PRS_ADHD_ and for 80 ICD-10 diagnoses after correction for multiple testing (Table 2, Figure S1, Table S3). Overall, the top five medical conditions with the strongest evidence for associations were chronic obstructive pulmonary disease (COPD) (OR=1.15, CI=1.11-1.18), obesity (OR=1.13, CI=1.11-1.15), type 2 diabetes (OR=1.11, CI=1.09-1.14), dorsalgia (OR=1.08, CI=1.07-1.10), and polyarthrosis (OR=1.09, CI=1.07-1.12).

**Table 2.**
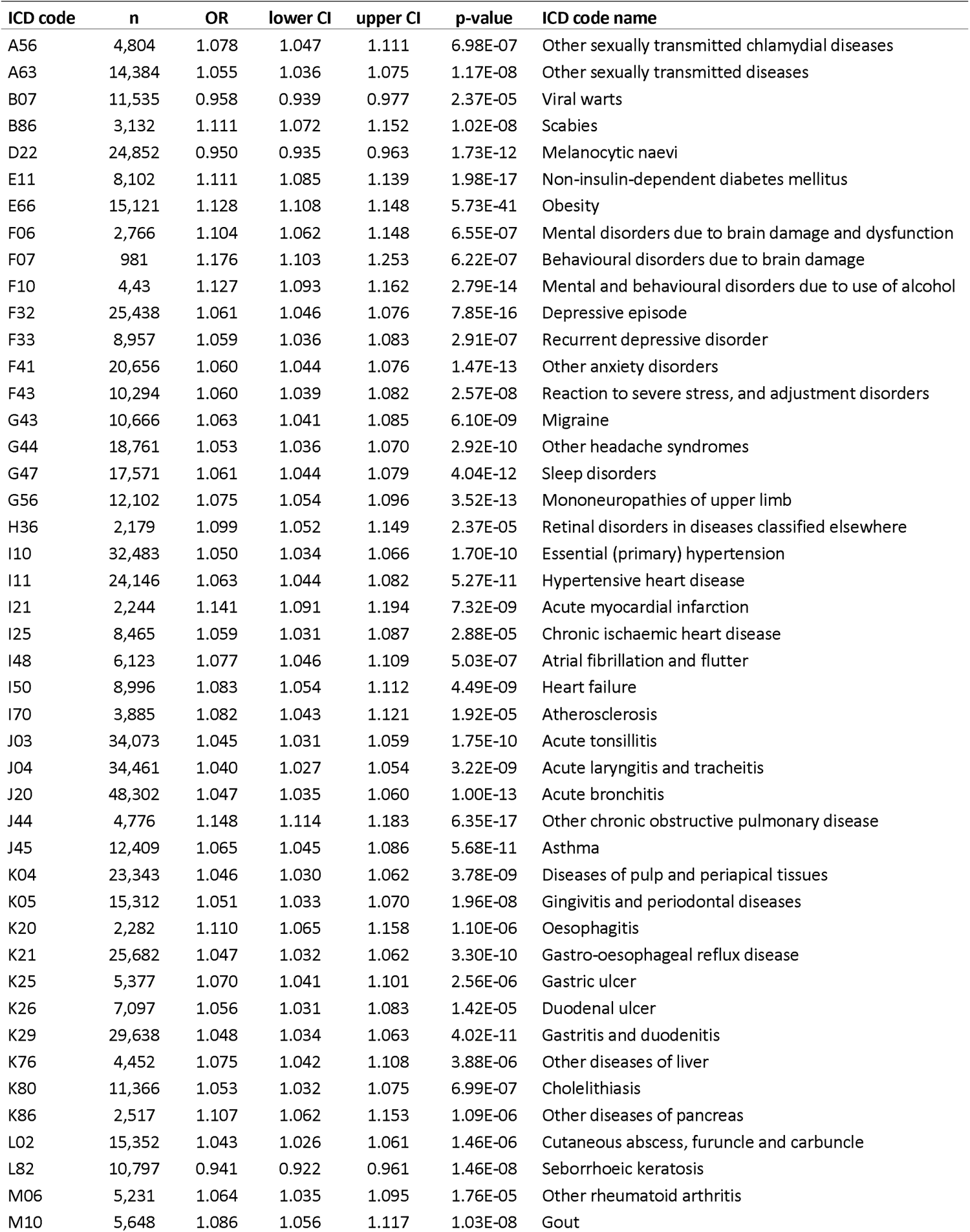
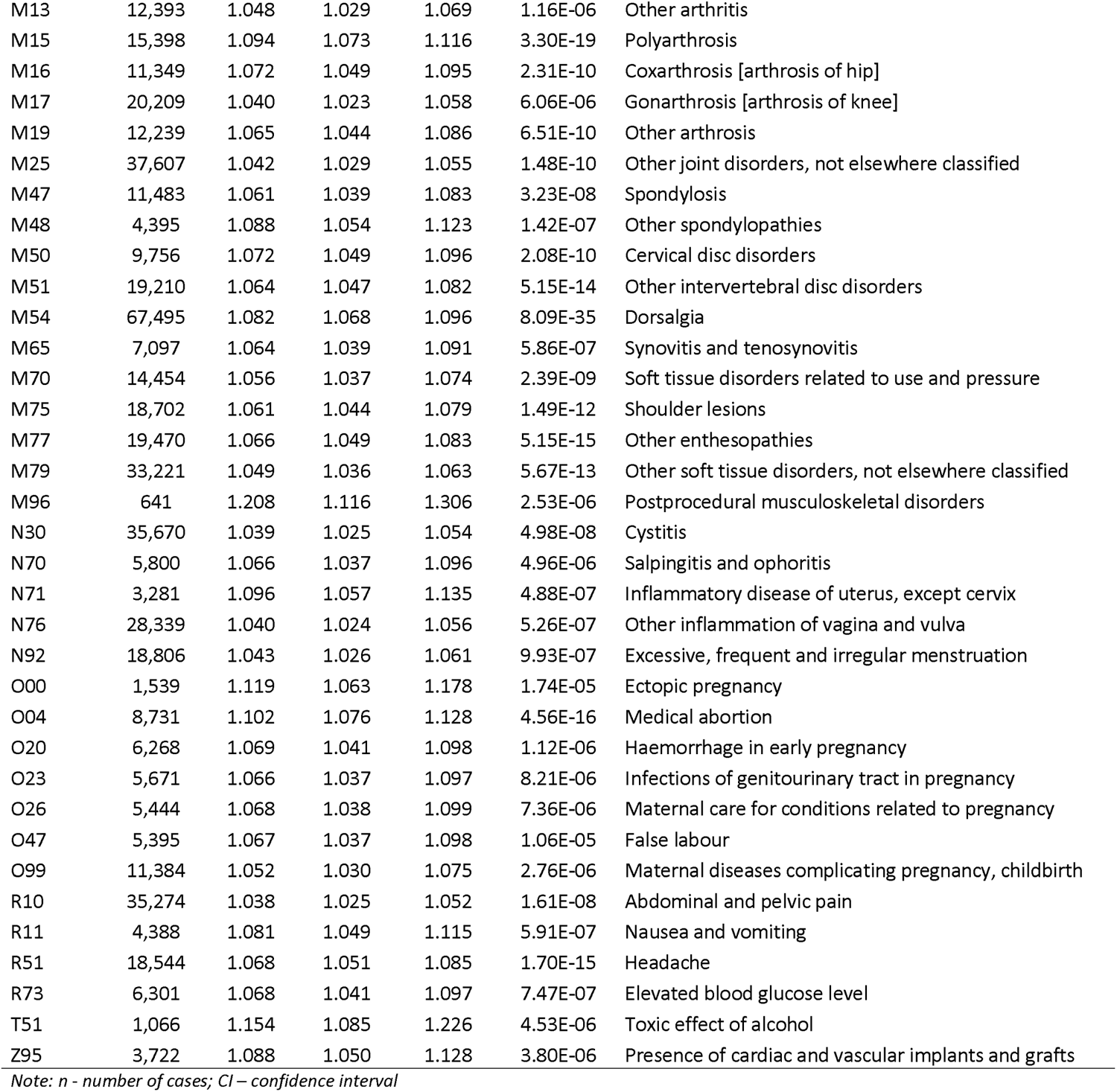
Associations between PRS_ADHD_ and ICD-10 codes after adjustment for multiple testing.

Sex-stratified analyses showed that for females, the top five associated medical conditions were obesity (OR=1.14, CI=1.11-1.16), polyarthrosis (OR=1.10, CI=1.08-1.13), medical abortion (OR=1.10, CI=1.08-1.13), dorsalgia (OR=1.09, CI=1.07-1.09), and headache (OR=1.08, CI=1.06-1.10). For males, the top five associated medical conditions were COPD (OR=1.17, CI=1.12-1.23), mental and behavioural disorders due to alcohol use (OR=1.11, CI=1.07-1.15), obesity (OR=1.11, CI=1.07-1.15), type 2 diabetes (OR=1.10, CI=1.06-1.14), and dorsalgia (OR=1.07, CI=1.04-1.09). Results are shown in Table S4 and Figures S2-S3.

The results in the full cohort (relatives not excluded) were largely similar, but additionally 52 associations passed correction for multiple testing potentially due to larger sample size and non-independence between observations (Table S5).

### PRS_ADHD_ deciles

The PRS_ADHD_ decile analysis of the 80 significantly associated medical conditions showed that individuals in the top PRS_ADHD_ decile had 70% higher risk for COPD (OR=1.70; CI=1.48-1.95); 59% higher risk for obesity (OR=1.59; CI=1.47-1.73), 51% higher risk for toxic effects of alcohol (OR=1.51; CI=1.16-1.97), 45% higher risk for type 2 diabetes (OR=1.45; CI=1.30-1.61), and 34% higher risk for polyarthrosis (OR=1.34, CI=1.23-1.47), compared to individuals in the lowest PRS_ADHD_ decile. The results using top vs middle PRS_ADHD_ decile were mostly in line with the results from top vs bottom PRS_ADHD_ decile analysis, but were non-significant for 23 medical conditions, e.g. other sexually transmitted diseases, heart failures, atherosclerosis, acute bronchitis, other arthritis and pregnancy related conditions (Figure 1; Tables S6-S7). The results comparing the high PRS_ADHD_ group separately in females and males are shown in Figure 2 and Tables S8-S9. 11 phenotypes were only available in females (such as pregnancy-related conditions). The majority of the associations in females and males were in the same direction. Of the phenotypes that were available in both gender, 15 associations were significant only in females (including, cystitis, cholelithiasis, gastro-oesophageal reflux disease (GORD), migraine, gastric ulcer, other arthrosis, other joint disorders, other spondylopathies, abdominal and pelvic pain and other sexually transmitted disease (STD) diagnoses). In contrast, associations with personality and behavioural disorders due to brain damage/dysfunction, toxic effects of alcohol and cardiac/vascular implants and grafts were only significantly associated in males.

**Figure 1.**
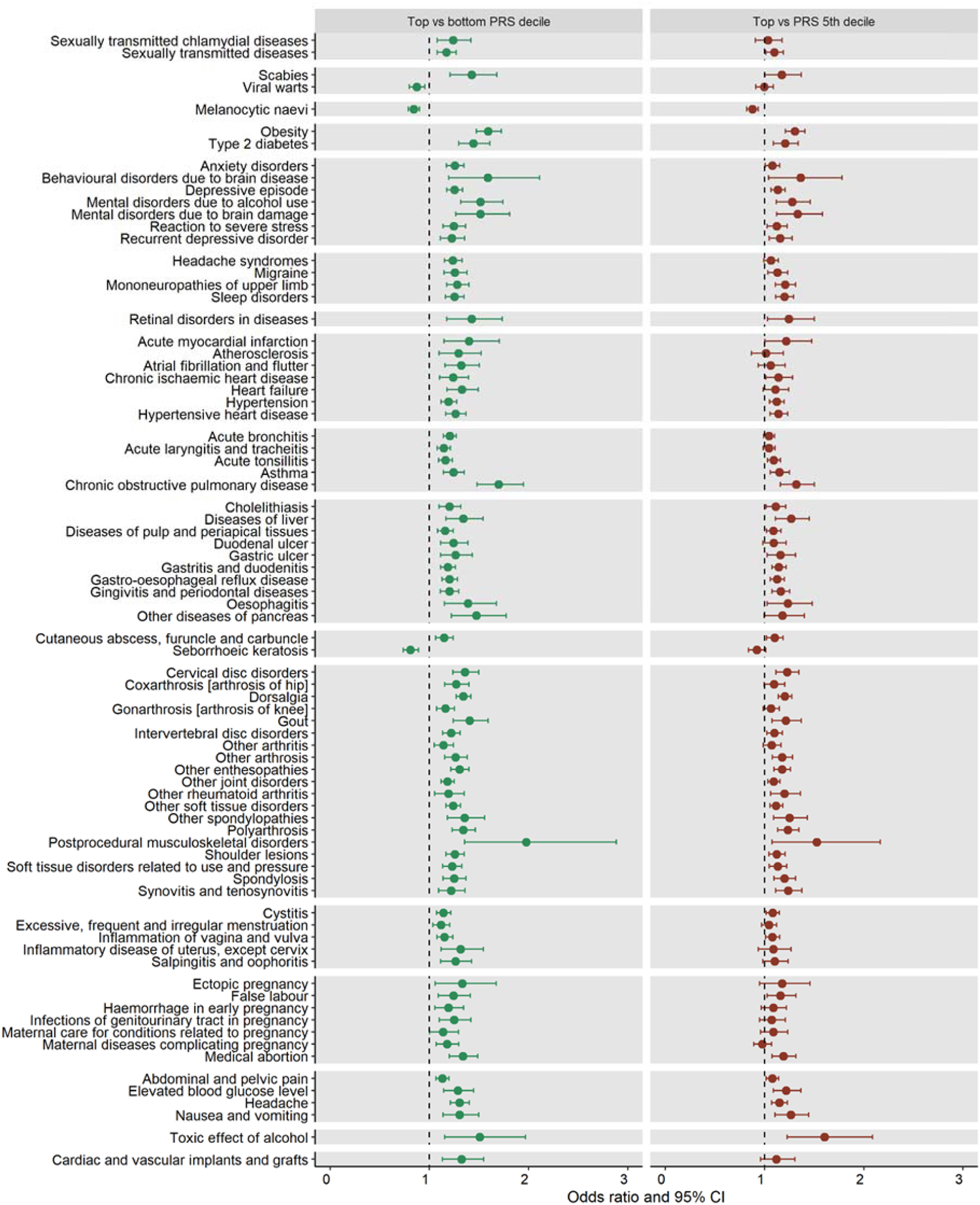
Comparison of associations between top vs bottom and top vs medium PRS_ADHD_ risk on ICD-10 codes after adjustment for multiple testing.

**Figure 2.**
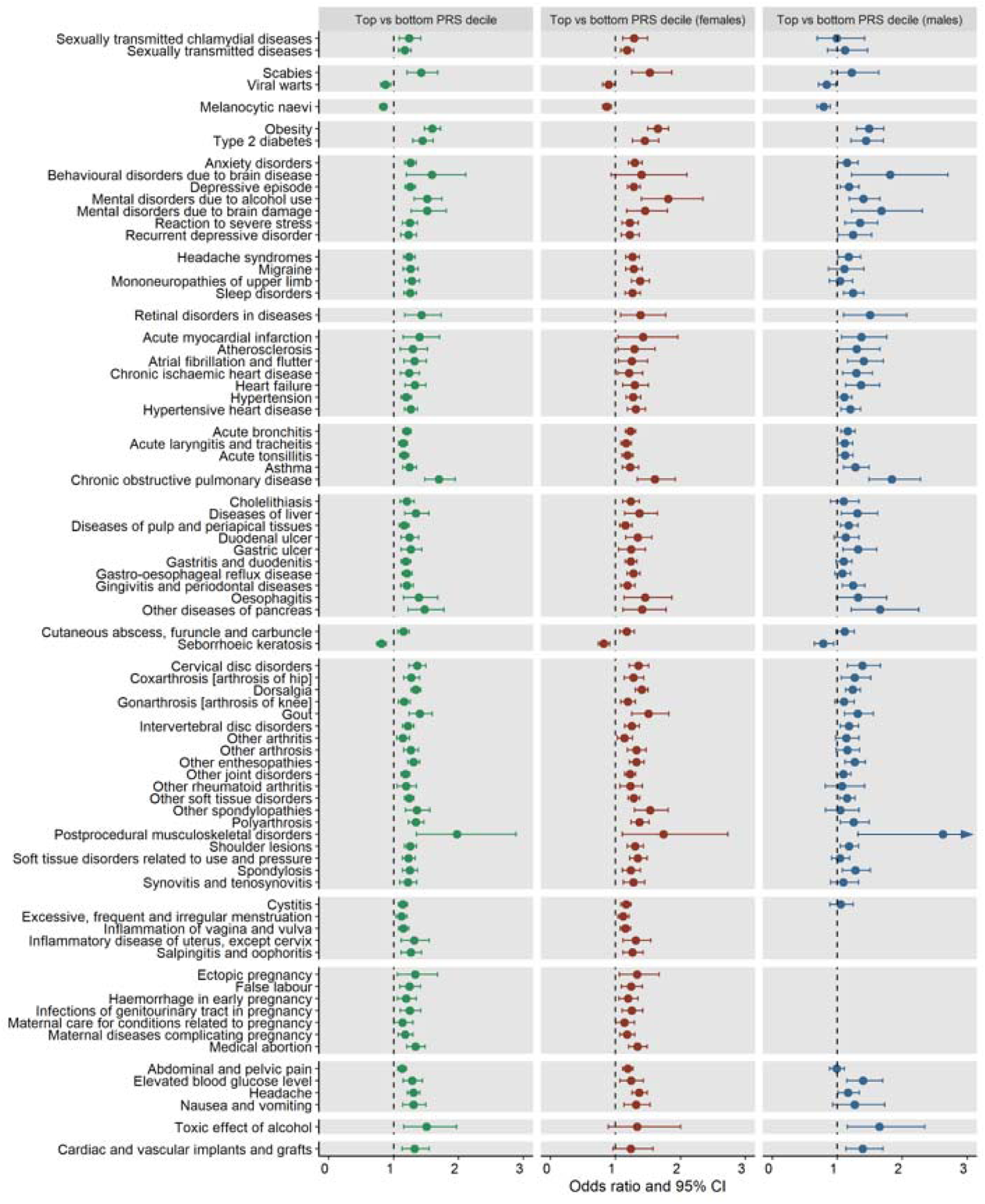
Comparison of associations between high PRS_ADHD_ risk and ICD-10 codes in females and males.

### Secondary analyses

#### Comparison of effects across PRS_ADHD_, ADHD diagnosis and questionnaire-based ADHD analyses

In the ADHD diagnosis analysis, 18 medical conditions had insufficient case numbers (<10 cases) to be included in these analyses (Table S10). We observed significant associations between ADHD diagnosis and 31 medical conditions, of which 6 medical conditions showed protective associations (e.g. other STDs, melanocytic nevus, excessive and irregular menstruation, medical abortion and pregnancy related conditions) (Figure 3). Overall, 40% of the PRS_ADHD_ associations were also observed in the ADHD diagnosis analysis. We observed the strongest evidence for associations with psychiatric disorders, e.g. behavioural disorders due to brain disease (OR=6.42, CI=4.08-10.11), reaction to severe stress (OR=4.70, CI=3.83-5.76), recurrent depressive disorders (OR=3.97, CI=3.08-5.11), but also with asthma (OR=2.11, CI=1.64-2.70) and acute bronchitis (OR=1.72, CI=1.43-2.06). In the questionnaire-based ADHD analysis, the direction of the effects were largely similar as in the PRS decile analysis and 78% of the PRS_ADHD_ associations were observed in the questionnaire-based ADHD analysis. We observed the strongest evidence for associations with recurrent depressive disorders (OR=3.78, CI=3.44-4.16), anxiety disorders (OR=2.31, CI=2.14-2.50), polyarthrosis (OR=1.94, CI=1.70-2.20), sleep disorders (OR=1.94, CI=1.77-2.13), and asthma (OR=1.64, CI=1.48-1.81). These results are shown in Figure 3, and Tables S10-S11.

**Figure 3.**
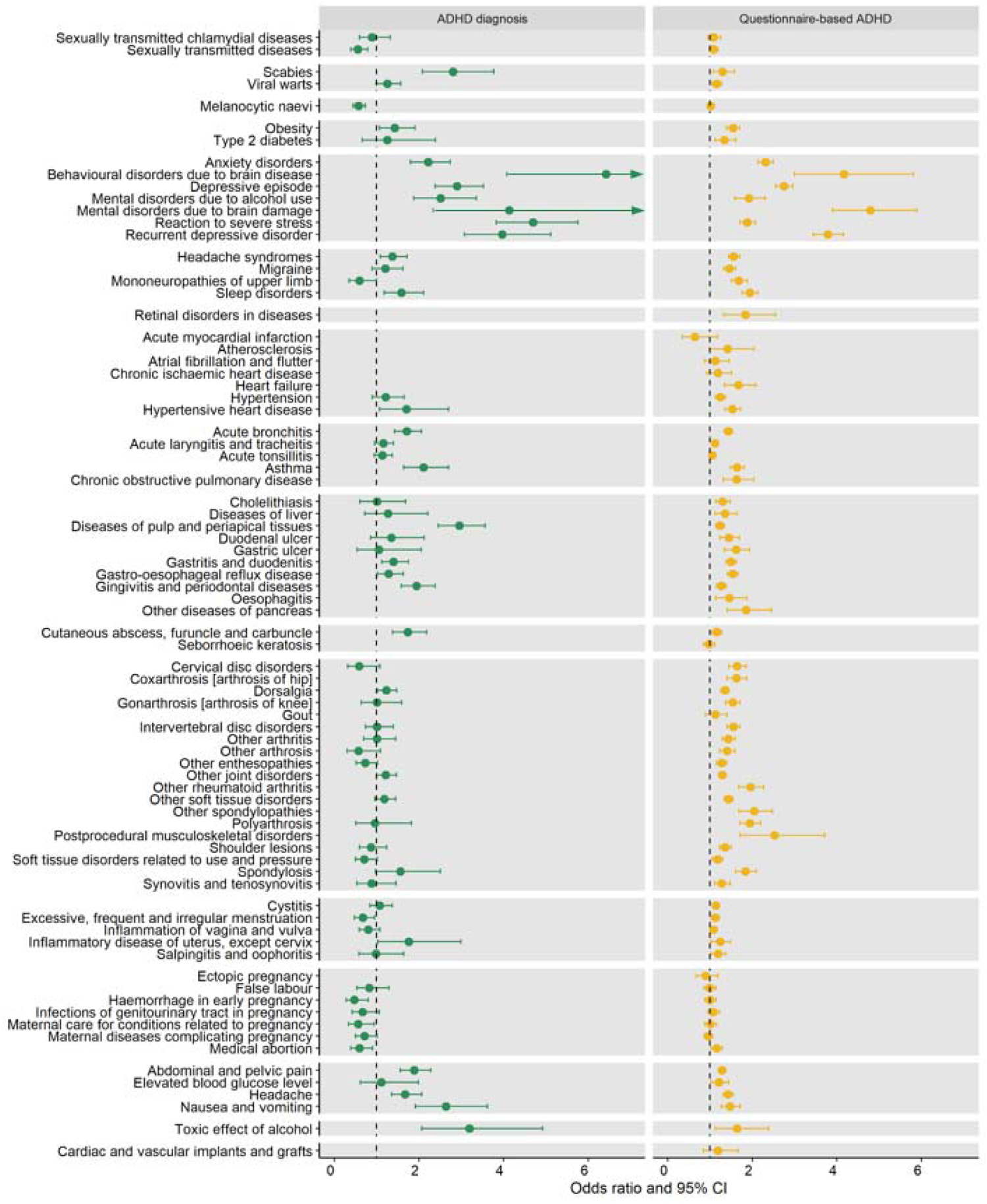
Comparison of associations between ADHD diagnosis and questionnaire-based ADHD on ICD-10 codes after adjustment for multiple testing.

### Causal mediation analysis

Our findings from causal mediation analysis showed that depression was a significant mediator for all 78 medical conditions (two depression diagnoses were excluded from the analysis), but the proportion of the effect between PRS_ADHD_ and medical conditions that was mediated by depression for majority of outcomes was generally small, ranging from 2 to 16%. Only for mental disorders, e.g. sleep disorders, anxiety disorders, reaction to severe stress and mental disorders due to brain damage/dysfunction, the mediation effect accounted for more than 20% of the observed associations between PRS_ADHD_ and respective ICD-10 diagnosis (Table S12).

## Discussion

In this study we performed a PheWAS between PRS_ADHD_ and EHR-based diagnoses across all ICD-10 categories, using nationwide, population representative clinical data with up to 17 years follow-up. This approach enabled the investigation of the associations between genetic liability to ADHD and medical conditions in a population without a history of ADHD diagnosis. Overall, our results indicate robust associations between ADHD genetic liability and 80 medical conditions, with obesity, dorsalgia, polyarthrosis, COPD and type 2 diabetes among the top associated medical conditions.

These results are in line with previous findings on the associations between ADHD diagnosis and comorbid medical conditions (Du Rietz et al., 2021; Momen et al., 2020). Furthermore, similarly to our findings, other studies using PRS_ADHD_ (Kember et al., 2021; Leppert et al., 2020) also showed associations between PRS_ADHD,_ mental and physical health problems, indicating a shared genetic background between ADHD and medical conditions. Another large-scale genetic study, based on UK Biobank data, found that genetic liability to multiple complex traits in adulthood, such as type 2 diabetes, obesity, peripheral artery disease and polyarthritis, increases the risk for ADHD in childhood, which further shows the role of underlying genetic effects between physical health and ADHD across development (García-Marín, Campos, Cuéllar-Partida, et al., 2021).

Several of the observed associations between PRS_ADHD_ and medical conditions in non-diagnosed individuals are consistent with the few previous genetically informative studies on diagnosed ADHD and medical comorbidities, such as the nervous system (Du Rietz et al., 2021), respiratory (Du Rietz et al., 2021; García-Marín, Campos, Kho, et al., 2021), musculoskeletal (Du Rietz et al., 2021) and cardiometabolic diseases (Du Rietz et al., 2021; Garcia-Argibay et al., 2022). Several studies have demonstrated that some of the observed associations between medical conditions and ADHD diagnosis were mediated by lifestyle factors, such as tobacco use and alcohol misuse (Faraone et al., 2021; Garcia-Argibay et al., 2022), which may reflect the impulsive behaviour characteristic to ADHD. Although we focused only on undiagnosed individuals in this study, the PRS_ADHD_ likely captures the genetic predisposition to ADHD related traits, such as impulsivity, which is strongly associated with risk behaviour and unhealthy lifestyle (Solmi et al., 2021). Therefore, future studies should explore whether health behaviour mediates the associations between ADHD genetic predisposition and medical conditions in different populations where detailed information on health behaviour is available. However, given the strong comorbidity between ADHD and depression, we utilized the diagnosis information to test mediation by depression diagnosis. As expected, we found that depression was a significant mediator for all observed medical conditions, but the effect from depression alone was rather small. Additionally, it is possible that adults with undiagnosed or subclinical ADHD may be more likely diagnosed with depression, either because of potentially express symptoms overlapping with depression (e.g. concentration problems, inattention) or being in an increased depression risk due to their unmanaged ADHD.

A recent Mendelian Randomization study showed a bidirectional causal effect between ADHD and obesity-related traits (e.g. waist-hip-ratio (WHR) and BMI-adjusted WHR) and reported genetic overlap between BMI and ADHD (Karhunen et al., 2021). It has been also shown that obesity increases the risk for a chronic inflammatory state (Andersen, Murphy, & Fernandez, 2016), which can lead to several diseases, such as hypertension, type 2 diabetes, cardiovascular disease, and musculoskeletal diseases (Furman et al., 2019; Nikiphorou & Fragoulis, 2018). There is also some evidence that inflammation plays an important role in the aetiology of ADHD (Dunn, Nigg, & Sullivan, 2019). It has been reported that individuals with ADHD had an increased inflammatory marker interleukin 6 (IL-6) which has been linked with increased risk of diabetes (Donfrancesco et al., 2020; Wang et al., 2013). Similarly, the role of obesity and inflammation markers are well described in the aetiology of cholelithiasis and GORD (El-Serag, 2008; Littlefield & Lenahan, 2019). Therefore, it is plausible that obesity can act as a mediator on the pathway between ADHD and several medical conditions. In fact, the study by Garcia-Argibay and colleagues found that BMI mediated associations between PRS_ADHD_ and medical conditions like hypertension, type 2 diabetes, migraine and sleep disorders (Garcia-Argibay et al., 2022).

The sex-stratified analysis generally showed similar associations in males and females. Several conditions that were statistically significant only in one gender (e.g. diseases of the musculoskeletal and digestive system, personality and behavioural disorders due to brain damage/dysfunction, toxic effects of alcohol and cardiac/vascular implants and grafts), we observed similar effect sizes in both genders, potentially indicating insufficient case numbers in the sex-stratified analyses. However, we also observed some sex differences. The association with sexually transmitted chlamydial diseases was more strongly associated in females. The literature supports the link between ADHD and risky sexual behaviour. Several studies have shown that ADHD is associated with higher risk of sexually transmitted diseases and more and younger age at first pregnancies (Hechtman et al., 2016; Hosain, Berenson, Tennen, Bauer, & Wu, 2012). The study by Leppert and colleagues (Leppert et al., 2020) also demonstrated the association between PRS_ADHD_ and younger age at first sexual intercourse in the UK Biobank. Although, the prevalence of chlamydia is similar in females and males, the disease is more often asymptomatic in males. Considering that women are more frequently screened in a routine gynaecological care, it is possible that the stronger association observed in females can be explained by higher likelihood of receiving a diagnosis (Dielissen, Teunissen, & Lagro-Janssen, 2013). Other associations observed more strongly in females were with cystitis, migraine, cholelithiasis, abdominal and pelvic pain and GORD diagnoses. Cystitis, migraine, abdominal and pelvic pain and cholelithiasis have been reported to be more common in women, which has been suggested to be explained by female hormones and inflammation processes (Allais et al., 2020; Curran, 2015; Littlefield & Lenahan, 2019; Patnaik et al., 2017). However, although GORD symptoms are equally prevalent in males and females (El-Serag, 2008), it has been suggested that oestrogen may mediate the association in females (Nilsson, Johnsen, Ye, Hveem, & Lagergren, 2003).

Although the majority of the associations observed in the PRS_ADHD_ analysis sample were in the same direction as in the ADHD diagnosis and questionnaire-based ADHD analysis samples, we observed opposite effects for some conditions in the ADHD diagnosis sample (such as conditions only expressed in females, e.g. pregnancy related conditions). It is possible that the protective associations observed with these conditions could be affected by treatment effects, potentially reducing the likelihood of psychosocial or environmental risk factors (Fuller-Thomson & Lewis, 2015) and altering reproductive behaviour (Østergaard, Dalsgaard, Faraone, Munk-Olsen, & Laursen, 2017; Skoglund et al., 2019). Additionally, it is possible that the diagnosed ADHD sample in EstBB is healthier, more educated and practicing healthy lifestyle compared to the average ADHD patient.

Our results suggest that ADHD-related medical conditions are also present in individuals without ADHD diagnosis but who have high genetic liability for ADHD. Evidence suggests that ADHD in adults is likely underdiagnosed and undertreated across countries (Fayyad et al., 2017). For example, ADHD lifetime prevalence in EstBB is 0.5% and yearly prevalence of ADHD was 0,8% in 2015-2020 in Estonia overall. This further highlights the importance of improving screening and management of ADHD in adult populations, in order to reduce the risk of severe physical comorbidities in individuals with subclinical or currently undiagnosed ADHD. This is also supported by our findings of the ADHD screening questionnaire, which revealed that 8,7% of the participants have high ADHD risk. However, for broader applicability, future studies should replicate these findings in other cohorts with varying rates of ADHD prevalence among adults.

### Strengths and limitations

The major strength of this study is inclusion of medical conditions across all ICD-10 diagnoses codes based on EHR with 17 years of follow-up, as well as combining genetic data with EHR and questionnaire data. EHR data is, as compared to self-reported data, not affected by recall bias and has better validity for many diagnoses as information is collected prospectively. Furthermore, we used data from a population-based biobank with a large sample size and high coverage of the whole population, thereby improving our statistical power to detect associations of even small effects.

However, this study also has some limitations. First, although EHR are a comprehensive data source, it only includes individuals with more severe symptoms and/or diseases who have received medical treatment. This may lead to misclassification bias, particularly in psychiatric categories, given the low EHR-based ADHD prevalence in EstBB. It is also possible that some individuals who received their ADHD diagnosis in childhood could have been left out from the study as EHR covers the period from 2004-2020. However, ADHD is often underdiagnosed in middle age and older adults because ADHD in adulthood has been recognized fairly recently (Franke et al., 2018). Availability of data from 2004 can also affect our mediation analysis results as we cannot be certain that depression was not present prior to any of the medical conditions identified in the PRS_ADHD_ analysis. Second, population-based cohort studies may be affected by selection bias as healthier and better educated people are more likely to participate in health studies (Larsson, 2021). The “healthy volunteer” selection bias has been previously described in the UK Biobank (Fry et al., 2017) and may also exist in the EstBB. Third, given the small number of ADHD cases in our sample, we could not test the effect of ADHD medications on the results. It is well reported that ADHD drug treatment can help to reduce various negative health outcomes and decrease risk of risky behaviour (Faraone et al., 2021). It is possible that the protective and statistically non-significant associations observed with some phenotypes in the ADHD diagnosis analysis could be affected by treatment effects. However, it is also plausible that individuals with ADHD diagnosis in the EstBB differ substantially from the average individual with ADHD diagnosis due to selection bias. Fourth, since the ADHD diagnosis analytical sample had higher mean birth year, the observed findings may have been affected by underestimation of associations as some diseases typically emerge in older age.

## Conclusion

Our study showed that genetic liability for ADHD is associated with increased risk for various medical conditions in individuals without ADHD diagnosis history. The results largely mirror the known associations between diagnosed ADHD and physical disease comorbidities. Although PheWAS design does not allow to conclude which specific mechanisms underly behind the observed associations, it provides insights for future studies to further investigate potential underlying pathways. Better knowledge about mechanisms explaining these associations can have a significant public health impact, as better detection and timely management of ADHD symptoms may help to reduce the risks for adverse medical conditions across the life span.

## Data Availability

According to the existing EU and Estonian data protection regulations, sensitive data (medical, genetic) cannot be made freely available. However, data access is possible following the standard data access procedure: https://genomics.ut.ee/en/content/estonian-biobank

## Acknowledgements

We thank all participants and staff of the Estonian biobank for their contribution to this research. The analytical work of EstBB was carried out in part in the High Performance Computing Center of the University of Tartu. We also thank Liisi Panov from the National Institute for Health Development for providing country-wide ADHD prevalence estimates and Silva Kasela for helping with causal mediation analysis.

Estonian Biobank Research Team: Andres Metspalu, Lili Milani, Tõnu Esko, Reedik Mägi, Mari Nelis and Georgi Hudjashov.

