## Supplementary material for "Associations between attention-deficit hyperactivity disorder genetic liability and ICD-10 medical conditions in adults: Utilizing electronic health records in a Phenome-Wide Association Study"

**Supplementary information**

**Supplementary methods**

Study population

The first set of 52,000 participants were recruited over 10-year period (2002-2011). The collection of samples and health data was set up consisting of GP’s and other medical personnel in private practices and hospitals or in the recruitment offices of the Estonian Genome Centre of University of Tartu (EGCUT) (1). The recruitment of next set of 150,000 participants started in 2018. Participants were recruited during promotion events, media campaigns, as well as through visiting GP´s and hospitals. Data collection was also conducted via questionnaire, which consists of 330 questions that gathered information about participant’s lifestyle, education, occupation, as well as anthropometric characteristics. However, through linkage with electronic health records and national registries, EstBB provides extensive follow-up data, which means that phenotype data can be periodically updated. The recruitment of participants is ongoing in a smaller scale and current available sample size is approximately 210,000.

Genotyping and imputation

All 200,000 EstBB participants have been genotyped using the Global Screening Array (GSA) from Illumina. The genotype calling for the GSA arrays was performed with the Illumina’s GenomeStudio V2010.3 software. The genotype calls for rare variants on the GSA array were corrected using the zCall software (version May 8th, 2012). After variant calling, the data was filtered using PLINK (v.1.90) by sample (call rate >95%, no sex mismatches between phenotype and genotype data, heterozygosity < mean +-3 SE) and marker (HWE p-value >1×10-6, call rate >95%, and additionally by Illumina GenomeStudio GenTrain score >0.6, Cluster Separation Score >0.4). Before the imputation, variants with MAF <1% and C/G or T/A polymorphisms, as well as indels were removed, as these genotype calls do not allow precise phasing and imputation (2). Prephasing was carried out using the Eagle v2.3 software (3) (number of conditioning haplotypes Eagle2 uses when phasing each sample was set to: --Kpbwt=20000) and imputation was executed using Beagle v.28Sep18.793 with an effective population size ne=20,000 (4,5). As a reference, the Estonian population specific imputation reference of 2297 WGS samples was used (6).

**Supplementary Figure 1. Associations between PRS_ADHD_ and ICD-10 codes**

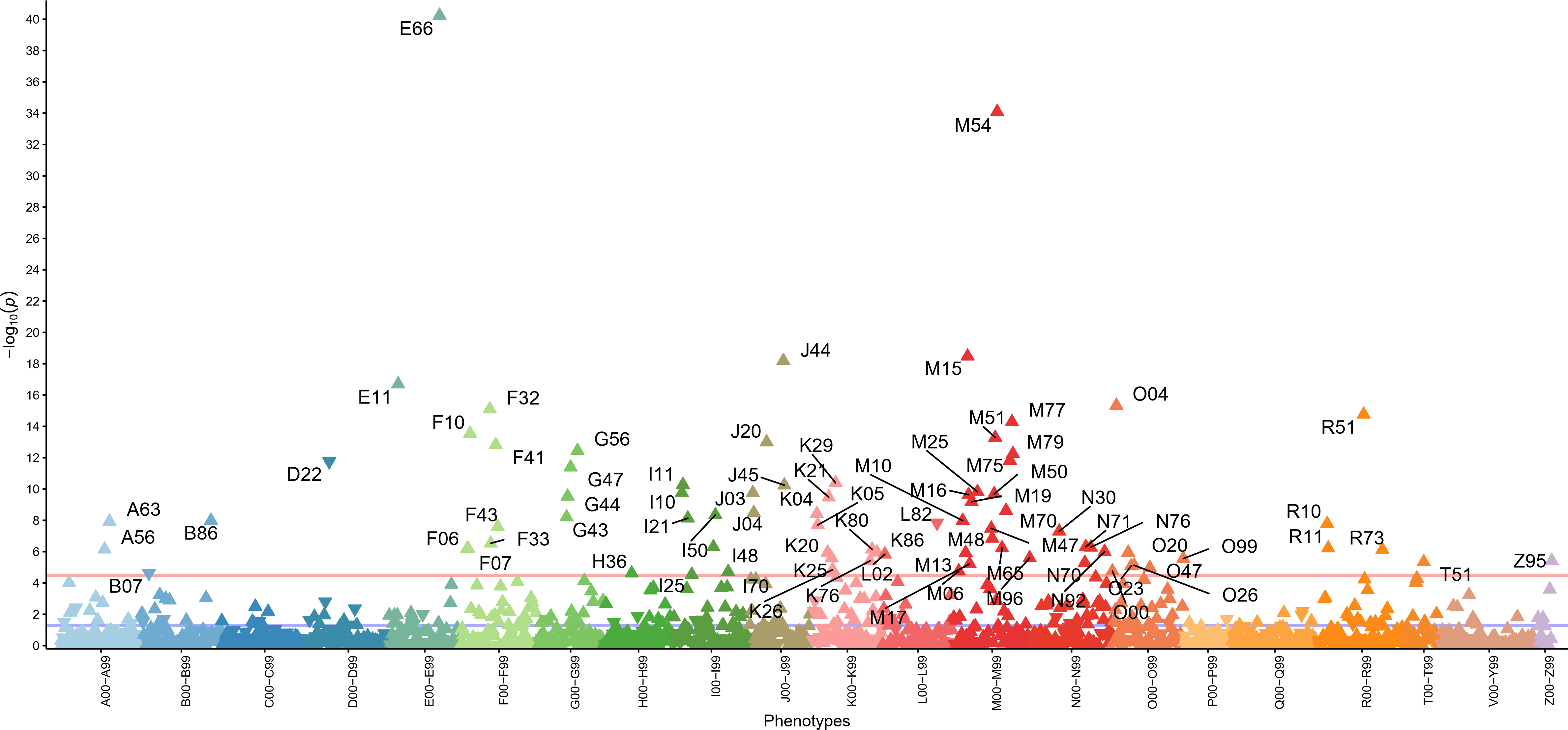

*Note: The X axis indicates groups of ICD10 main codes coloured respectively and Y axis −log10 of the p values. Each triangle in the plot represents one ICD-10 main code and the direction of the triangle represents direction of effect. Red line— Bonferroni-corrected significance level (3.3x10^-5^). Phenotypes passed the Bonferroni correction from the lowest to highest p-values: E66-obesity; M54-dorsalgia;M15-polyarthrosis; J44-chronic obstructive pulmonary diseases; E11-non-insulin dependent diabetes; O04-medical abortion; F32-depressive episode; R51-headache; M77-other enthesopathies; F10-mental and behavioural disorders due to use of alcohol; M51-other intervertebral disc disorders; J20-acute bronchitis; F41-other anxiety disorders; G56-mononeuropathies of upper limb; M79-other soft tisuse disorders; M75-shoulder lesion; D22-melanocytic naevi; G47-sleep disorders; K29-gastritis and duodenitis; I11-hypertensive heart disease; J45-asthma; M25-other joint disorders; I10-primary hypertension; J03-acute tonsillitis; M50-cervical disc disorders; M16-coxarthrosis; G44-other headache syndromes; K21-gastro-oesophageal reflux disease; M19-other arthrosis; M70-soft tissue disorders related to use, overuse and pressure; J04-acute laryngitis and tracheitis; K04-diseases of pulp and periapical tissues; I50-heart failure; G43-migraine; I21-acute myocardial infarction; B86-scabies; M10-gout; A63-other predominantly sexually transmitted diseases; L82-seborrhoeic keratosis; R10-abdonminal and pelvic pain; K05-gingivitis and periodontal diseases; F43-reaction to severe stress, M47-spondylosis; N30-cystitis; M48-other spondylopathies; F33-recurrent depressive disorder; N71-inflammatory disease of uterus; I48-atrial inflammation of vagina and vulva; M65-synovitis and tenosynovitis; R11-nausea and vomiting; F07-personality and behavioural disorders due to brain damage; F06-other mental disorders due to brain damage; A56-other sexually transmitted chlamydial diseases; K80-cholelithiasis; R73-elevated blood glucose level; N92-excessive, frequent and irregulaar menstruation; K86-other diseases of pancreas; K20-oesophagitis; O20-haemorrhage in early pregnancy; M13-other arthritis; L02-cutaneous abscess, furuncle and carbuncle; M96-postprocedural musculoskeletal disorders; K25-gastric ulcer; O99-other maternal diseases; Z95-presence of cardiac and vascular implants and grafts; K76-other diseases of liver; T51-toxic effect of alcohol; N70-salpingitis and oophoritis; M17-gonarthrosis; O26-maternal care for other conditions related to pregnancy; O23-infections of genitourinary tract in pregnancy; O47-false labour; K26-duodenal ulcer; O00-ectopic pregnancy; M06-other rheumatoid arthritis; I70-atherosclerosis; H36-retinal disorders in diseases classified elsewhere; B07-viral warts; I25-chronic ischaemic heart disease*

**Supplementary Figure 2. Associations between PRS_ADHD_ and ICD-10 codes in women**

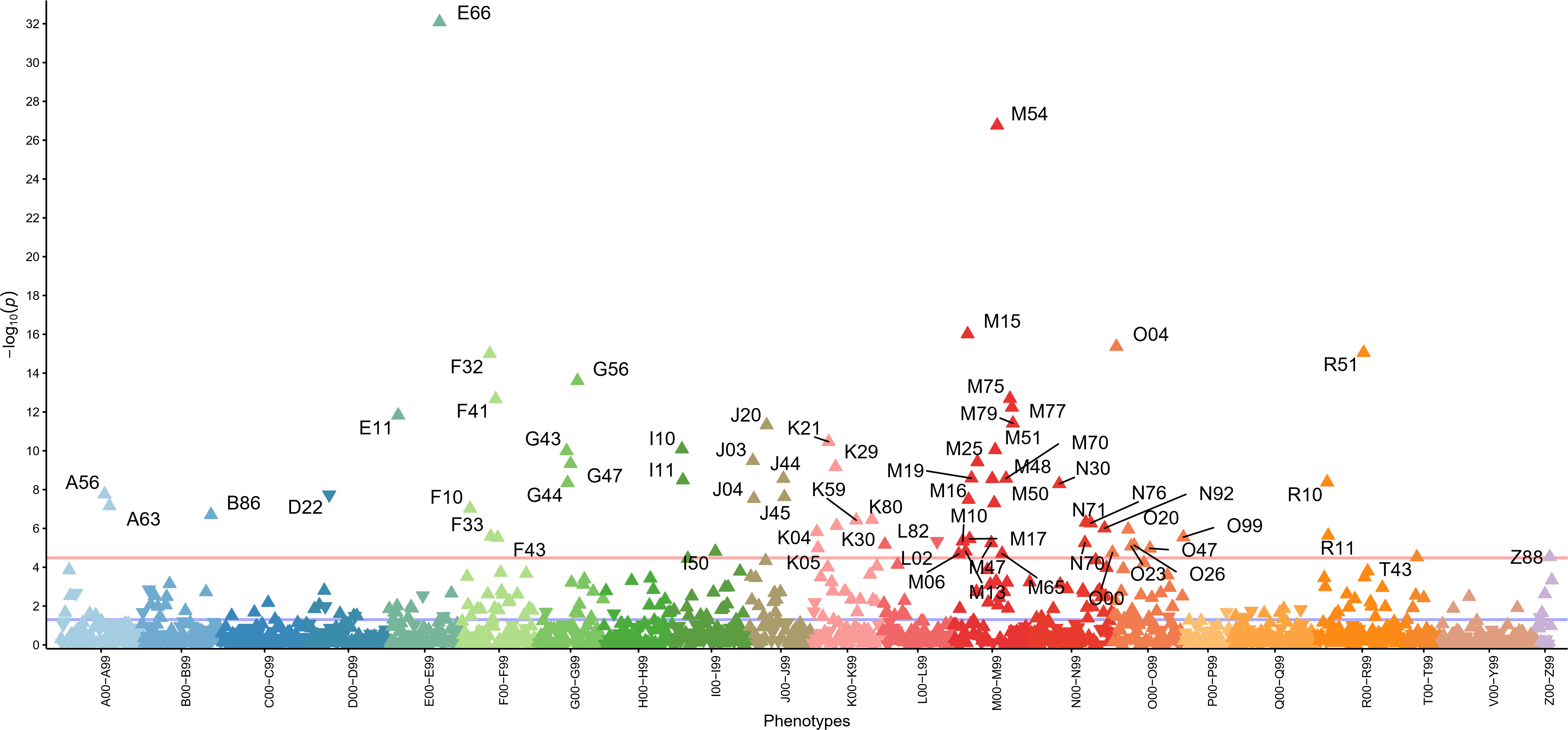

*Note: The X axis indicates groups of ICD10 maincodes coloured respectively and Y axis −log10 of the p values. Each triangle in the plot represents one ICD10 main code and the direction of the triangle represents direction of effect. Red line— Bonferroni-corrected significance level (3.3x10^-5^). Phenotypes passed the Bonferroni correction: E66-Obesity; M54-Dorsalgia; M15-Polyarthrosis; O04-Medical abortion; R51-Headache; F32-Depressive episoode; G56-Mononeuropathies of upper limb; M75-Shoulder lesions; F41-Other anxiety disorders; M77-Other enthesopathies; E11-Non-insulin-dependent diabetes mellitus; M79-Other soft tissue disorders, not elsewhere classified; J20-Acute bronchitis; K21-Gastro-oesophageal reflux disease; I10-Essential (primary) hypertension; M51-Other intervertebral disc disorders; G43-Migraine; J03-Acute tonsillitis; M25-Other joint disorders, not elsewhere classified; G47-Sleep disorders; K29-Gastritis and duodenitis; M19-Other arthrosis; M70-Soft tissue disorders related to use, overuse and pressure; J44-Other chronic obstructive pulmonary disease; M48-Other spondylopathies; I11-Hypertensive heart disease; R10-Abdominal and pelvic pain; G44-Other headache syndromes; N30-Cystitis; A56-Other sexually transmitted chlamydial diseases; D22-Melanocytic naevi; J45-Asthma; J04-Acute laryngitis and tracheitis; M16-Coxarthrosis [arthrosis of hip]; M50-Cervical disc disorders; A63-Other predominantly sexually transmitted diseases, not elsewhere classified; F10-Mental and behavioural disorders due to use of alcohol; B86-Scabies; K80-Cholelithiasis; K59-Other functional intestinal disorders; N71-Inflammatory disease of uterus, except cervix; N76-Other inflammation of vagina and vulva; K30-Dyspepsia; N92-Excessive, frequent and irregular menstruation; O2-Haemorrhage in early pregnancy; K04-Diseases of pulp and periapical tissues; R11-Nausea and vomiting; F33-Recurrent depressive disorder; O99-Other maternal diseases classifiable elsewhere but complicating pregnancy, childbirth and the puerperium; F43-Reaction to severe stress, and adjustment disorders; M17-Gonarthrosis [arthrosis of knee]; M10-Gout; L82-Seborrhoeic keratosis; N70-Salpingitis and oophoritis; M47-Spondylosis; L02-Cutaneous abscess, furuncle and carbuncle; O26-Maternal care for other conditions predominantly related to pregnancy; O23-Infections of genitourinary tract in pregnancy; K05-Gingivitis and periodontal diseases; O47-False labour; M13-Other arthritis; I50-Heart failure; O00-Ectopic pregnancy; M6-Synovitis and tenosynovitis; M06-Other rheumatoid arthritis; Z88-Personal history of allergy to drugs, medicaments and biological substances; T43-Poisoning by psychotropic drugs, not elsewhere classified*

**Supplementary Figure 3. Associations between PRS_ADHD_ and ICD-10 codes in men**

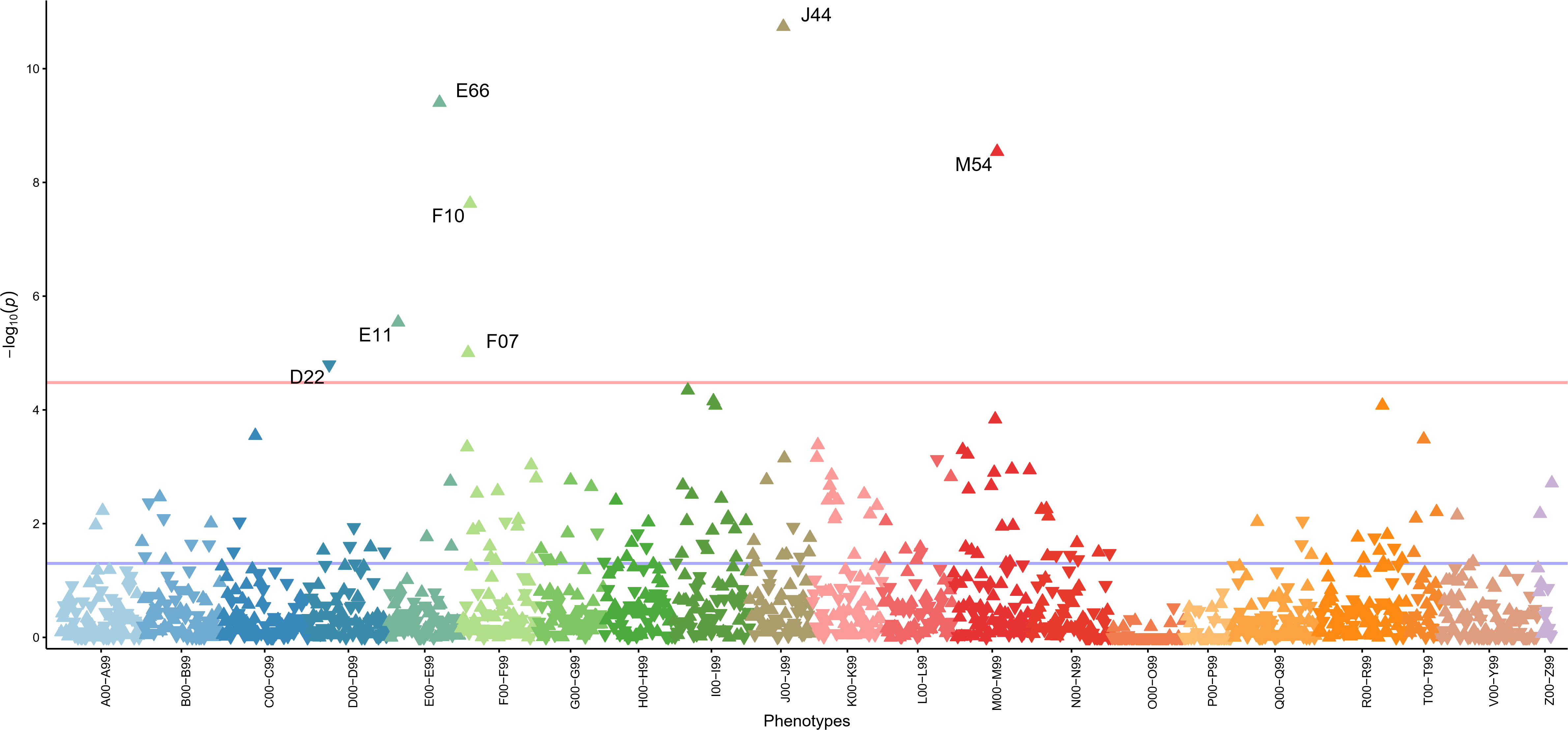

*Note: The X axis indicates groups of ICD10 maincodes coloured respectively and Y axis −log10 of the p values. Each triangle in the plot represents one ICD10 main code and the direction of the triangle represents direction of effect. Red line— Bonferroni-corrected significance level (3.3x10^-5^). Phenotypes passed the Bonferroni correction: J44-Other chronic obstructive pulmonary disease; E66-Obesity; M54-Dorsalgia; F10-Mental and behavioural disorders due to use of alcohol; E11-Non-insulin-dependent diabetes mellitus; D22-Melanocytic naevi*
